# REFINE: Closing the Loop Between Large Language Models and Symbolic Rules in Clinical NLP

**DOI:** 10.64898/2026.08.11.26360118

**Authors:** Nan Wang, Alexandra Kakadiaris, Chenyu Li, Rongrong Wang, Jaerong Ahn, Yashan Wang, Sunyang Fu

**Affiliations:** Dell Medical School, The University of Texas at Austin, Austin, TX, USA; Center for TEAM-AI, McWilliams School of Biomedical Informatics Houston, TX, USA; Department of Biomedical Informatics, University of Pittsburgh School of Medicine Pittsburgh, PA, USA

**Keywords:** clinical natural language processing, large language models, symbolic NLP, electronic health records, clinical information extraction

## Abstract

Symbolic clinical natural language processing (NLP) systems remain widely used for extracting clinical concepts from electronic health record (EHR) narratives, but maintaining rule resources requires extensive manual error analysis and rule refinement. This study investigates whether large language models (LLMs) can assist in identifying extraction errors and generating candidate rules to improve symbolic clinical NLP systems. Using error reports derived from a multi-site evaluation of a previously validated symbolic model for cognitive and neuropsychiatric-related clinical concepts, we developed a human-in-the-loop framework, REFINE. The framework first uses LLMs to classify extraction errors and generate explanatory reasoning, which can then be incorporated into prompts for rule generation. Three LLMs (GPT-5.2, GPT-4o, GPT-4o-mini) were evaluated under four prompting conditions. LLM-generated rule sets improved performance compared with the baseline NLP-CAM system, increasing F1-score from 0.37 to 0.58. These findings suggest that LLMs can support scalable rule refinement for symbolic clinical NLP systems.

## I. Introduction

Clinical artificial intelligence is increasingly moving toward hybrid architectures that combine the expressive power of large language models (LLM) with the precision and auditability of symbolic reasoning, a paradigm often termed neuro-symbolic AI [1]. In this context, symbolic refers to representations built from explicit, human-readable constructs such as logical expressions, lexical patterns, and structured knowledge representations, rather than the distributed numerical weights that underlie neural models [2]. A rule is the core construct: a discrete, explicitly authored mapping (e.g., a regular-expression pattern, lexical trigger, or logical condition) that specifies how a given linguistic pattern should be interpreted or classified. Rules are grounded in lexicons, curated term sets and synonym lists that supply the domain vocabulary from which patterns are constructed. Because each of these constructs is authored and read in human interpretable form, every decision the system makes, from the concept it extracts to the reasoning behind a correction, can be traced back to an explicit symbolic expression.

Within this landscape, symbolic clinical natural language processing (NLP) can provide the structured, verifiable knowledge layer upon which trustworthy clinical AI systems are built. Rather than treating symbolic systems as a legacy alternative to neural approaches, emerging frameworks recognize symbolic NLP as the interpretable core that anchors AI-driven clinical workflows in transparent, clinically validated logic [3]. This distinction is especially consequential as healthcare moves toward clinical Digital Twins (DT), defined as dynamic, patient-level computational representations that continuously integrate structured and unstructured EHR data to simulate disease trajectories and support individualized clinical decision-making [4]. Because digital twins rely on explicit, mechanistic representations of patient states and trajectories, the concept extraction pipelines feeding them needed to be auditable, controllable, and institutionally portable [5], [6].

With high-quality annotation data and rigorous lexicon and pattern development and validation processes, symbolic approaches have demonstrated strong performance across a wide range of clinical NLP benchmarks [7], [8], [9], including top-ranked submissions in the 2014 i2b2/UTHealth deidentification challenge [10], [11] and the 2009 i2b2 medication extraction challenge [12], establishing their reliability in high-stakes information extraction settings. By encoding clinical knowledge directly through curated vocabularies and regular-expression rules, symbolic systems extract clinically meaningful concepts from narrative text while maintaining full traceability of every extraction decision [5], [13], [14]. In addition, symbolic resources can be easily modified and transferred across institutions, which is an essential component in multi-site research networks. For example, within the Evolve to Next-Gen Accrual to Clinical Trials Network (ENACT) national research network, collaboratively developed NLP algorithms have been deployed across participating institutions to standardize concept extraction from clinical narratives and integrate results into a shared common data model [15].

Scalability is another important consideration in the deployment of clinical NLP. Large healthcare institutions have increasingly emphasized platform-based architectures that combine symbolic NLP engines with enterprise clinical data pipelines and scalable computing infrastructure. For example, Mayo Clinic recently designed an NLP platform to support high-throughput processing of unstructured clinical narratives across enterprise-scale EHR systems by integrating symbolic extraction components with distributed data processing workflows [16]. These architectures enable symbolic systems to operate efficiently on large volumes of clinical documentation while maintaining the interpretability of symbolic methods. of unstructured clinical narratives across enterprise-scale EHR systems by integrating symbolic extraction components with a distributed data processing workflow [16]. These architectures enable symbolic NLP systems to operate efficiently on large volumes of clinical documentation while maintaining the interpretability of symbolic methods.

Despite these advantages, symbolic NLP systems remain sensitive to linguistic variation and documentation heterogeneity. Clinical narratives frequently contain ambiguous terminology, abbreviations, contextual qualifiers, and institution-specific documentation styles. In addition, contextual modifiers such as negation, temporality, and experiencer can alter the interpretation of clinical concepts mentioned in text. Symbolic algorithms such as ConText use lexical triggers and contextual patterns to determine these attributes and improve the interpretation of extracted concept [17]. As a result, symbolic extraction systems often generate false positives (FP) and false negatives (FN) when applied across heterogeneous clinical environments. Identifying the underlying causes of these errors and updating rule resources accordingly typically requires extensive manual analysis and expert review. Error analysis is therefore a critical step in maintaining and improving symbolic clinical NLP systems [18]. However, systematic interpretation of extraction errors and consistent lexicon and rule refinement can be difficult to standardize across datasets and institutions. In multisite studies, differences in documentation practices, annotation standards, and error-reporting formats further complicate manual analysis of extraction failures [19], [20].

Recent advances in large language models (LLMs) have demonstrated strong capabilities in structured reasoning, information extraction, and natural language understanding tasks [21], [22]. Prior research has shown that LLMs can support a variety of biomedical text-mining applications, including literature analysis [23], [24], clinical information extraction [25], and interpretation of medical narratives [26]. These capabilities suggest that LLMs may also assist in analyzing errors in NLP systems and in supporting the refinement of symbolic extraction systems.

In this study, we developed a human-in-the-loop opensource framework, REFINE (Rule Error Feedback for Iterative Neural Editing), that leverages LLMs to assist in analyzing and correcting extraction errors in a symbolic clinical NLP system. Using error reports derived from a prior multi-site evaluation of an NLP-CAM extraction system [27], we designed a pipeline in which LLMs first interpret extraction errors and then generate candidate regularexpression rules intended to address those errors. The framework was implemented and evaluated in a real-world multisite study conducted as part of the ENACT NLP Working Group [28]. We evaluated three LLMs across multiple prompting conditions to determine whether incorporating LLM-generated error interpretations improves rule-generation performance.

## II. Methods

The proposed REFINE framework consists of four major stages. First, real-world extraction errors from a previously evaluated symbolic NLP system [28] were collected and harmonized into a structured dataset. Second, LLMs were used to analyze errors in the training dataset by assigning error classes and generating explanations of the likely causes of each failure. Third, the LLMs were prompted to generate candidate regular expression rules to address the identified errors. Finally, the generated rules were incorporated into the NLP-CAM rule resource files and evaluated to assess differences in concept-extraction performance.

### A. Real-World Error Construction

This study used error reports derived from a prior multisite evaluation of the NLP-CAM symbolic clinical NLP system, designed to extract cognitive and neuropsychiatric symptoms (CNS) from EHR notes [27] within the ENACT NLP Working Group Framework, which focuses on algorithms for identifying delirium-related clinical concepts. The ENACT consortium supports collaborative development and validation of NLP algorithms using shared data models and multi-site evaluation frameworks to improve the generalizability of clinical text-mining systems across healthcare environments [28].

The original evaluation included clinical data from four institutions representing distinct EHR environments: UTPhysicians (UTP), Memorial Hermann Health System (MHHS), Harris County Psychiatric Center (HCPC), and Beth Israel Deaconess Medical Center (BIDMC). In the parent study, NLP-CAM outputs were compared with manually annotated gold-standard labels generated through expert review. The extraction task focused on identifying delirium-related clinical concepts documented in clinical notes, including symptoms and descriptors associated with cognitive and neuropsychiatric symptoms (e.g., agitation, confusion, disorientation). Differences between system predictions and gold-standard annotations were recorded as extraction errors. For the present study, the unit of analysis was the extracted error report rather than the original clinical note.

Site-level error reports were provided as spreadsheets containing instances of false positives (FP) and false negatives (FN) produced by the NLPCAM system. False positives occur when the system predicts a concept absent from the gold standard, whereas false negatives occur when a gold-standard concept is not detected by the system. Each error record was associated with a norm, representing the predefined clinical concept category targeted by the extraction system (e.g., agitation, delirium, confusion). Norms correspond to the concept labels used within the NLP-CAM symbolic extraction framework [27]. Because the format and completeness of error reports varied across sites, preprocessing was required before the data could be integrated into a unified analysis pipeline. Some institutions provided complete error sentences and gold-standard labels, whereas others provided only triggering terms or omitted contextual fields required for downstream rule generation and evaluation. Records lacking essential information, such as the full sentence or gold standard concept label, were excluded.

After cleaning and harmonization, a unified dataset consisting of 754 error instances was constructed. Retained variables included UniqueID, site name, gold-standard norm, NLP-CAM norm, error type (FP/FN), a binary gold standard indicator (GS), and the corresponding sentence. The GS indicator was defined as 1 when the gold-standard norm corresponded to any clinical concept other than “none,” and 0 when the gold-standard norm was “none.” The dataset was then partitioned into training and test sets using a 50/50 stratified split by norm, ensuring that the distribution of clinical concept labels was preserved across both datasets.

### B. LLM Implementation and Error Classification

In the first stage, the training error dataset was provided to an LLM to assign structured error classes (common symbolic NLP failures such as negation, hypothetical statements) and generate explanatory rationales for each error instance. A set of candidate error classes was derived from our previously developed clinical NLP error taxonomy [18] and adapted for our study. Three LLMs were evaluated: GPT-5.2, GPT-4o, and GPT-4o-mini, accessed through the HIPAA-compliant Azure OpenAI platform.

The explanatory reasoning statement was optionally incorporated into the downstream rule-generation prompts for each error instance. For example, when processing the sentence “Neurological exam today is relatively nonfocal, but dizziness does improve with breath holding,” the models provided different interpretations of the underlying error:

- GPT-4o-mini – Absence of Context: The term “dizziness” is mentioned, but the sentence lacks sufficient clinical context to determine the significance of the finding.
- GPT-4o – Medical Evaluation: The sentence reflects a general neurological assessment rather than a definitive statement about the patient’s status, which may lead to this error classification.
- GPT-5.2 – Exclusion: The sentence describes dizziness and a nonfocal neurological exam and does not relate to impaired connectedness or communication; therefore, the predicted concept is unrelated to the clinical content.

Prompts consisted of a fixed system prompt describing the error-analysis task and a user prompt containing the sentence, norm, and error type. Model outputs were constrained to a structured JSON format containing the predicted error class and reasoning. All experiments were performed using deterministic decoding with a temperature of 0 to reduce response variability and improve reproducibility. Prompts were identical across models to ensure comparability.

### C. LLM-Assisted Rule Refinement

In the second stage, the LLM was prompted to generate regular expression rules to correct the identified extraction errors. These rules are pattern-matching expressions used by NLP-CAM to detect clinical concepts in clinical text. To evaluate the effect of different levels of error context information on rule generation, four prompting conditions were designed.

- Experiment 0 (E0) served as the baseline condition and did not include any generated error class or reasoning.
- Experiment 1 (E1) incorporated the LLM-predicted error class in addition to the baseline inputs.
- Experiment 2 (E2) included the LLM-generated reasoning but excluded the error class.
- Experiment 3 (E3) included both the predicted error class and the corresponding reasoning.

These conditions enabled a systematic evaluation of whether progressively richer contextual information derived from LLM-based error analysis improved rule-generation performance. For each model (GPT-4o-mini, GPT-4o, and GPT-5.2), rule generation was performed under all four experimental conditions, resulting in distinct rule sets for E0– E3. The generated rule sets fully replaced the original manually curated NLP-CAM rules, allowing evaluation to isolate the performance of LLM-generated rules without influence from the manually curated baseline rule set.

Each generated rule set was integrated into the NLP-CAM pipeline, and the system was re-executed on the held-out test dataset. Pipeline outputs were then aligned with the harmonized evaluation dataset. For each experiment, two fields were appended to the evaluation dataset: a predicted norm and a binary prediction indicator, where 1 indicates that at least one concept was extracted and 0 indicates no detection. Using the binary gold-standard indicator (GS), Classification outcomes were defined as follows: a True Positive (TP) occurs when the gold standard (GS) equals 1 and the prediction equals 1; a True Negative (TN) occurs when GS equals 0 and the prediction equals 0; a False Positive (FP) occurs when GS equals 0 and the prediction equals 1; and a False Negative (FN) occurs when GS equals 1 and the prediction equals 0. Performance metrics were then computed by aggregating counts across norms.

### D. User Interface Application Development

REFINE was implemented as a single-page web application backed by a lightweight Python Flask server running on localhost. The server exposed a REST API for reading and writing MedTagger rule files, executing shell scripts via subprocess, and returning annotation output for reevaluation. Rule file discovery was performed dynamically by parsing used_resources.txt and resources_rules_matchrules.txt, allowing the system to operate over arbitrary task configurations without modification. LLM-assisted rule suggestions were generated by querying a locally hosted language model through the Ollama inference server, using structured prompts that incorporated error context, annotation guidelines, and the current rule file contents (https://github.com/OHNLP/REFINE). Fig. 1 displays screenshots of the user interface.

**Fig. 1.**
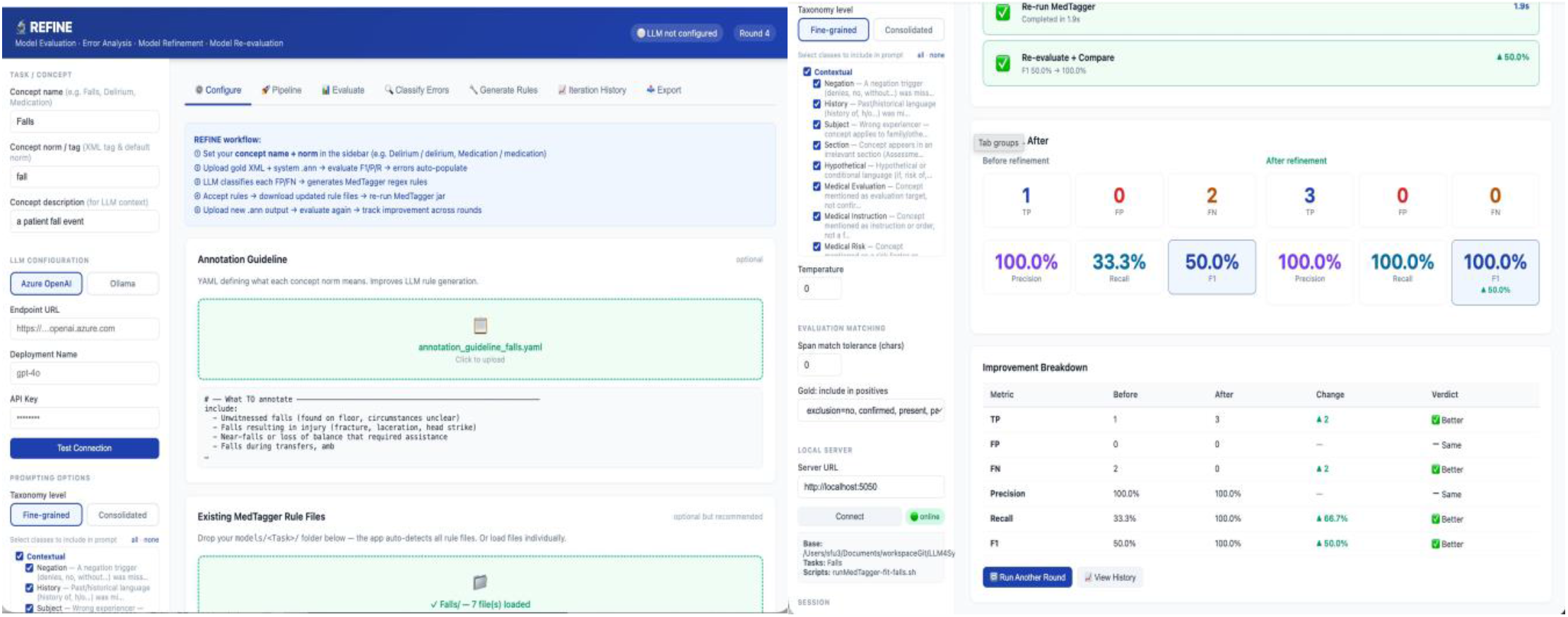
REFINE User Interface.

### E. Evaluation

Performance was evaluated on the test dataset using standard classification metrics derived from TP, TN, FP, and FN counts. Metrics reported include precision, recall (sensitivity), specificity, accuracy, NPV, and F1-score. Performance was summarized by norm for the baseline NLPCAM system and for each LLM-generated rule set. Additional analyses were stratified by site to examine potential variation in performance across institutions.

The primary objective of the evaluation was to determine whether LLM-generated rules could feasibly reduce extraction errors in a symbolic clinical NLP system. A secondary objective was to evaluate whether incorporating LLM-generated error classes and reasoning improves rule generation performance relative to prompting without intermediate error interpretation.

## III. Results

### A. Dataset Characteristics

After preprocessing and harmonization, the final dataset contained 754 extraction error instances, consisting of 560 false negatives (FN) and 194 false positives (FP). False negatives represent instances in which the system failed to detect a gold-standard concept, whereas false positives represent cases in which the system predicted a concept that was not present in the gold-standard annotation. Fig. 2 summarizes the distribution of error instances across clinical norms. False negative errors were most frequent for agitation (n = 109) and hallucination (n = 70), while several norms, such as disconnected and confusion, exhibited substantial numbers of both false positive and false negative errors. This distribution indicates that many clinically relevant concepts were under-detected by the baseline system, particularly those expressed through diverse lexical or contextual variations in clinical narratives.

**Fig. 2.**
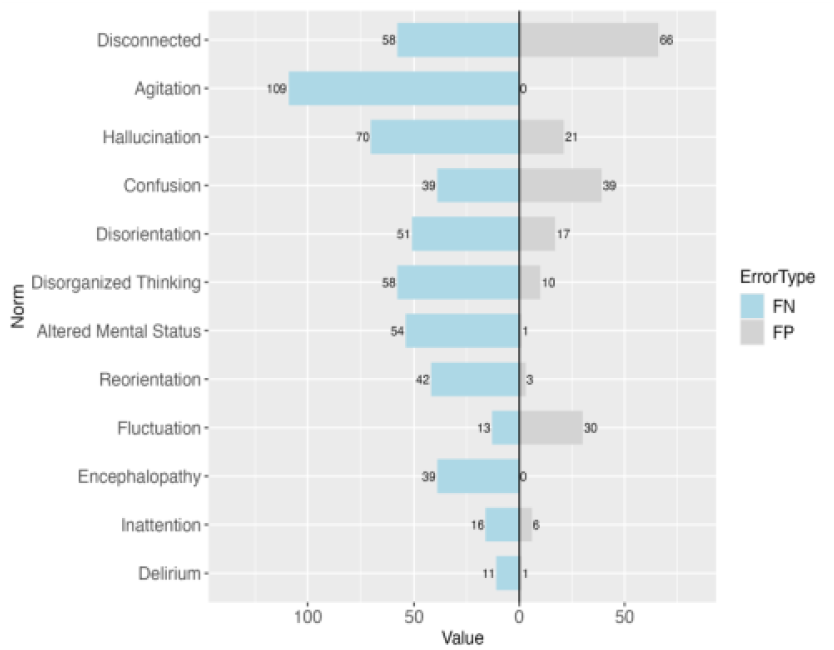
False Negative and False Positive Distribution.

### B. LLM Error Classification Outputs

Error classification was performed on the training set to generate structured explanations of the causes of symbolic extraction failures. A representative example illustrating differences in error classification across models can be found in the git repository (https://github.com/OHNLP/REFINE). Although the models analyzed the same sentence, they attributed the error to different causes, reflecting variation in how contextual and semantic information were interpreted. Fig. 2 showcases the overall distribution of predicted error classes. Across models, structural error categories accounted for a large proportion of classifications. GPT-4o-mini frequently classifies sentences as the absence of context, whereas implied inference and exclusion were more commonly assigned by GPT-4o and GPT-5.2. Contextual errors, such as negation and history, were also commonly detected. In contrast, lexical error categories appeared less frequently overall, although GPT-5.2 identified a notable number of synonym-related errors.

### C. Rule Generation Outputs

In the second stage of the pipeline, we prompted the LLMs to generate regular expression rules intended to correct the extraction errors identified during the error classification stage. Across experiments, generated rules frequently introduced lexical expansions and morphological variants of target concepts. For example, several rules included pattern variants such as agitated, agitation, and restlessness. Larger models generally produced more comprehensive pattern expansions, while smaller models sometimes generated simpler or narrower patterns. In a small number of cases, the generated regular expression patterns required minor syntactic correction before being incorporated into the NLP-CAM rule files. These corrections primarily involved resolving malformed regular expression syntax. Such adjustments were infrequent for GPT-5.2 and GPT-4o, but more frequent for GPT-4o-mini.

### D. Performance by Model

Fig. 3 summarizes system performance across all models baseline system. Among the evaluated models, GPT-5.2 achieved the strongest overall results, with its best performance observed in Experiment 2 (F1 = 0.58). GPT-4o demonstrated moderate improvements, reaching a maximum F1-score of 0.50 under Experiment 2. GPT-4o-mini produced smaller improvements overall, with its best performance observed in Experiment 3 (F1 = 0.45). The best-performing experiments and metrics are highlighted in bold in Table 5. Overall, Experiment 2 produced the strongest performance across models.

**Fig. 3.**
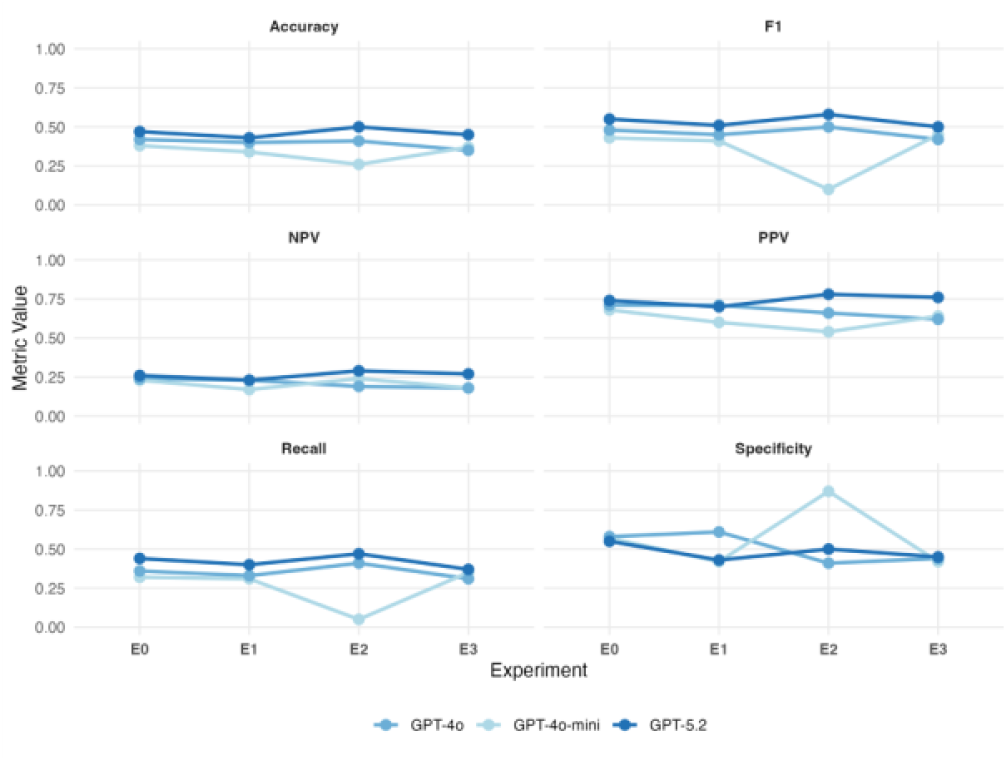
Performance metrics across experiments and LLM models.

### E. Performance by Error Class and Model

Fig. 4 compares model performance stratified by error class and model. The best-performing configurations, GPT-4o-mini (E3), GPT-4o (E2), and GPT5.2 (E2), were used for comparison. Overall, GPT-5.2 achieved the strongest performance, with consistent improvements observed as model capability increased. F1score of 0.50 under Experiment 2. GPT-4o-mini produced smaller improvements overall, with its best performance observed in Experiment 3 (F1 = 0.45). Overall, Experiment 2 produced the strongest performance across models. Performance also varied across norms. Concepts with more explicit terminology, such as agitation and delirium, generally achieved higher F1-scores, whereas concepts requiring greater contextual interpretation showed lower performance. Site-level comparisons show a similar trend: performance generally increases as model capability increases. However, performance at UTP was consistently lower than that of the other participating institutions.

**Fig. 4.**
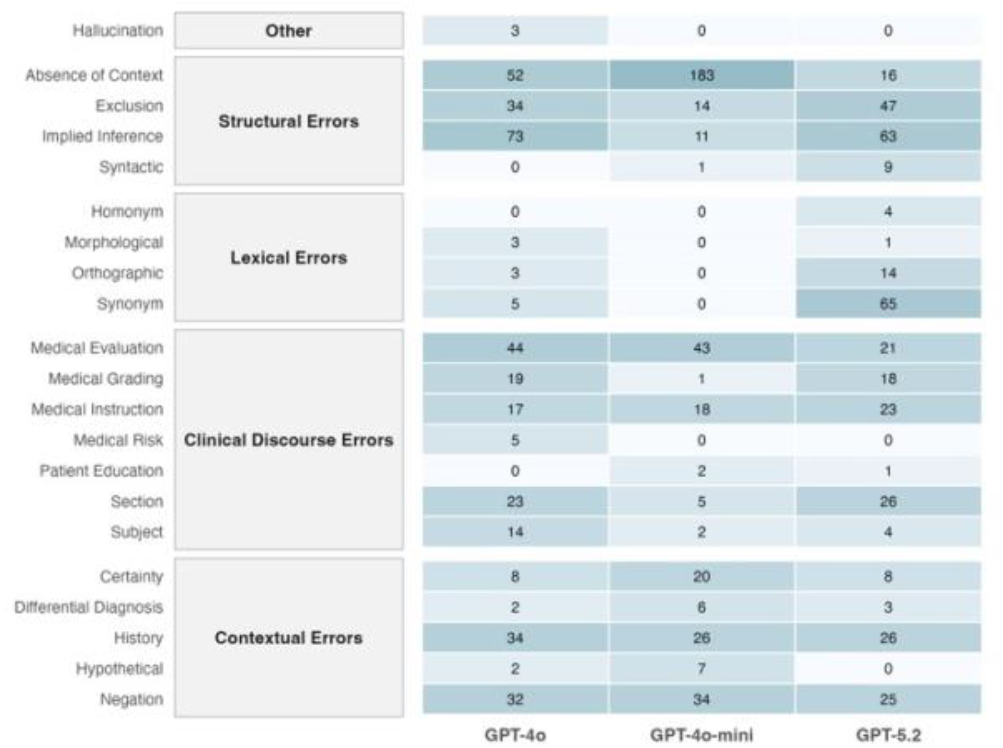
Heatmap of error counts by error class and LLM.

### F. Performance by Norm and Site

Fig. 5 compares model performance stratified by clinical norm and participating sites. The best-performing configurations, GPT-4o-mini (E3), GPT-4o (E2), and GPT5.2 (E2), were used for comparison. Overall, GPT-5.2 achieved the strongest performance, with consistent improvements observed as model capability increased. Performance also varied across norms. Concepts with more explicit terminology, such as agitation and delirium, generally achieved higher F1-scores, whereas concepts requiring greater contextual interpretation showed lower performance. Site-level comparisons show a similar trend in performance, generally increasing as model capability increases. However, performance at UTP was consistently lower than that of the other participating institutions.

**Fig. 5.**
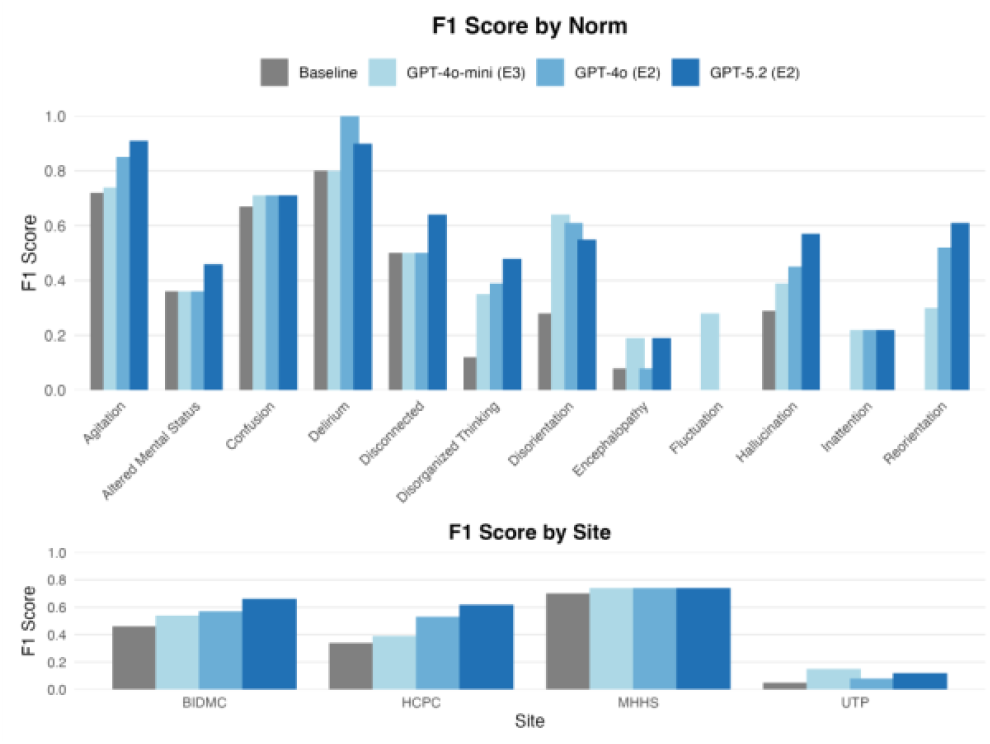
F1 score comparison per LLM across norms and clinical site.

## IV. Discussion

To address the challenges of manual error analysis and rule refinement in symbolic clinical NLP systems, we developed the REFINE framework, a human-in-the-loop pipeline that assists with interpreting extraction errors and generating candidate regular expression rules. Across all experiments, LLM-generated rule sets improved performance compared with the baseline NLP-CAM system. The baseline model achieved an F1 score of 0.37, whereas multiple LLM-generated configurations substantially increased performance. The best overall result was achieved by GPT-5.2 under Experiment 2, which reached an F1-score of 0.58, representing an approximate 57% improvement over the baseline system. GPT-4o achieved its best performance under Experiment 2 with an F1-score of 0.50, corresponding to a 35% improvement, while GPT-4o-mini achieved its best performance under Experiment 3 with an F1-score of 0.45, representing a 22% improvement over baseline. Across models, larger models generally produced stronger results. GPT-5.2 consistently outperformed GPT-4o and GPT-4omini across most experimental conditions, suggesting that model capability influences the quality and generalizability of generated rule patterns. The error class distribution also provides insight into the types of extraction failures encountered during rule generation. Structural errors, particularly the absence of context and implied inference, represented the largest proportion of errors across models, while contextual errors such as negation and history were also frequently observed. These patterns indicate that many extraction failures stem from difficulties interpreting contextual information in clinical narratives rather than from simple lexical mismatches. Such contextual dependencies are often challenging for symbolic systems because they require interpretation of broader sentence-level meaning rather than isolated keyword patterns.

### A. Interpretation of Results

Several patterns emerged across the experimental conditions. Experiments that incorporated LLM-generated reasoning (E2 and E3) generally produced stronger performance than prompts that did not include reasoning. For GPT-4o and GPT-5.2, Experiment 2—where the prompt included reasoning without explicit error class labels— produced the best overall results. One possible explanation is that reasoning provides richer contextual information about the underlying cause of extraction failures while still allowing the model flexibility to generate generalized rules. In contrast, GPT-4o-mini performed best in Experiment 3, which included both the error class and reasoning. For smaller models, the additional structured guidance provided by the error class label may help constrain the rule-generation process and compensate for more limited reasoning capability. Across models, precision (PPV) remained consistently higher than negative predictive value (NPV), suggesting that the generated rules were more effective at identifying true concept mentions than at correctly identifying the absence of concepts. This pattern indicates that while LLM-generated rules improved detection of positive cases, the systems still struggled to capture negative cases, likely due to contextual complexity and linguistic variability in clinical documentation.

Performance also varied across clinical norms and participating institutions. Differences across norms likely reflect variation in how cognitive and neuropsychiatric symptoms are expressed in clinical documentation. Concepts such as delirium and agitation are often described using explicit terminology and therefore achieved relatively high performance. In contrast, concepts such as disorganized thinking, fluctuation, and encephalopathy often require interpretation within a broader clinical context and may be expressed in diverse narrative descriptions. These contextual and linguistic variations make such concepts more difficult for symbolic systems to capture using pattern-matching rules alone. Performance differences were also observed across sites. In particular, the UTP site demonstrated substantially lower performance relative to other institutions. One possible explanation is variation in the distribution of clinical norms within the UTP dataset, which contained a higher proportion of more context-dependent symptoms such as encephalopathy, fluctuation, and hallucination. Additionally, institutional differences in clinical documentation practices may influence symbolic extraction performance, as narrative style, terminology usage, and note structure can vary across healthcare systems. However, additional studies incorporating larger samples and detailed analysis of site-specific documentation patterns would be required to confirm the underlying causes of these differences.

### B. Practical Implications

The findings of this study suggest several practical implications for the development of clinical NLP systems. First, LLMs may provide a scalable method for assisting with rule maintenance in symbolic extraction systems. By automatically generating candidate rules based on observed extraction errors, LLM-assisted workflows could substantially reduce the time required for manual rule refinement. Second, the results highlight the practical value of hybrid architectures that combine LLMs with symbolic systems. While LLMs demonstrate strong capabilities in interpreting contextual information and generating candidate rules, they are computationally expensive and resource-intensive for large-scale deployment. In contrast, symbolic systems are computationally efficient and well-suited for high-throughput processing but often require extensive manual effort and may struggle with context-dependent language. Integrating LLM-assisted rule generation with symbolic extraction pipelines may therefore leverage the strengths of both approaches. Finally, incorporating LLM-based reasoning into rule development workflows may improve the consistency and reproducibility of error analysis processes, which are often difficult to standardize in multi-site studies.

### C. Limitations

Several limitations should be considered when interpreting these results. First, the dataset consisted only of extraction errors derived from a prior evaluation study rather than the full corpus of clinical notes. As a result, the evaluation focused specifically on error correction rather than overall concept extraction performance. Second, the dataset was relatively small and exhibited imbalances across clinical norms and sites, which may have influenced performance estimates and contributed to variability in model results. Third, the generated rule sets entirely replaced the original NLPCAM rules. In practice, a hybrid approach combining manually curated rules with LLM-generated rules may provide greater stability and robustness. Finally, while the results demonstrate improvements in extraction metrics, the generated rules were not manually validated by clinical domain experts.

### D. Future Work

Future research could explore several directions to extend this work. One promising approach is to integrate LLM-generated rules into an iterative, human-in-the-loop workflow in which domain experts review and refine candidate rules generated by the model. Such an approach may combine the efficiency of automated rule generation with the expertise of clinical annotators. Another important direction is investigating site-level performance variation. Future studies using larger datasets could examine how differences in documentation practices, narrative structure, and terminology usage across institutions influence symbolic extraction performance. Understanding these site-specific characteristics may help improve the portability of symbolic NLP systems across healthcare environments. Finally, future studies could evaluate LLM-assisted rule generation on larger datasets and across additional clinical NLP tasks to assess the approach’s broader generalizability.

## IV. Conclusion

To address the challenges of manual error analysis and rule refinement in symbolic clinical NLP systems, we developed a framework, REFINE, and applied it to a multisite ENACT case study. Using error reports from a prior evaluation of the NLP-CAM system, we implemented a pipeline in which LLMs first interpret extraction errors and then generate candidate regular-expression rules to address them. Across multiple experimental conditions, LLM-generated rule sets improved extraction performance relative to the baseline NLP-CAM system, with the best-performing configuration achieving an F1-score of 0.58 compared with 0.37 for the baseline system. These results demonstrate the potential of LLMs to support scalable rule refinement for symbolic clinical NLP pipelines. Rather than replacing symbolic systems, LLMs may serve as complementary components that assist with error interpretation and rule development. By reducing the manual effort required for rule maintenance, LLM-assisted workflows may improve the scalability and adaptability of symbolic clinical NLP systems across heterogeneous healthcare environments.

## Data Availability

The data used in this study are private and institutionally restricted and are therefore not publicly available.

